# Blood Cholesterol and Triglycerides Associate with Right Ventricular Function in Pulmonary Hypertension

**DOI:** 10.1101/2024.01.20.24301498

**Authors:** Battoul Fakhry, Laura Peterson, Suzy A.A. Comhair, Jacqueline Sharp, Margaret M. Park, W.H. Wilson Tang, Donald R. Neumann, Frank P. DiFilippo, Samar Farha, Serpil C. Erzurum, Anny Mulya

**Author notes:** Corresponding author: Anny Mulya, PhD, Lerner Research Institute, Cleveland Clinic, 9500 Euclid Avenue, Cleveland, Ohio 44195. deceased (Donald R. Neumann). “This work has not been published previously and it is not under consideration for publication elsewhere. All authors gave approval for this manuscript to be published.”.

## Abstract

**Background:** Blood lipids are dysregulated in pulmonary hypertension (PH). Lower high-density lipoproteins cholesterol (HDL-C) and low-density lipoproteins cholesterol (LDL-C) are associated with disease severity and death in PH. Right ventricle (RV) dysfunction and failure are the major determinants of morbidity and mortality in PH. This study aims to test the hypothesis that dyslipidemia is associated with RV dysfunction in PH.

**Methods:** We enrolled healthy control subjects (n=12) and individuals with PH (n=30) (age: 18-65 years old). Clinical characteristics, echocardiogram, 2-[18F] fluoro-2-deoxy-D-glucose positron emission tomography (PET) scan, blood lipids, including total cholesterol (TC), triglycerides (TG), lipoproteins (LDL-C and HDL-C), and N-terminal pro-B type Natriuretic Peptide (NT-proBNP) were determined.

**Results:** Individuals with PH had lower HDL-C [PH, 41±12; control, 56±16 mg/dL, *p*<0.01] and higher TG to HDL-C ratio [PH, 3.6±3.1; control, 2.2±2.2, p<0.01] as compared to controls. TC, TG, and LDL-C were similar between PH and controls. Lower TC and TG were associated with worse RV function measured by RV strain (*R*=–0.43, *p*=0.02 and *R=*–0.37, *p*=0.05 respectively), RV fractional area change (*R*=0.51, *p*<0.01 and *R*=0.48, *p*<0.01 respectively), RV end-systolic area (*R*=–0.63, *p*<0.001 and *R*=–0.48, *p*<0.01 respectively), RV end-diastolic area: *R*=–0.58, *p*<0.001 and *R*=–0.41, *p*=0.03 respectively), and RV glucose uptake by PET (*R*=–0.46, *p*=0.01 and *R*=–0.30, *p*=0.10 respectively). NT-proBNP was negatively correlated with TC (*R*=*–*0.61, *p*=0.01) and TG (*R*=–0.62, *p*<0.02) in PH.

**Conclusion:** These findings confirm dyslipidemia is associated with worse right ventricular function in PH.

## Introduction

Pulmonary hypertension (PH) is characterized by pulmonary vascular remodeling and increased pulmonary arterial pressure that lead to right ventricle (RV) dysfunction and ultimately failure^1^. Despite advances in therapies, patients with PH still face severe long-term morbidity and mortality^2^, and a reduction in quality of life^3^. The prognosis of PH, irrespective of its etiology, depends on the ability of the RV to adapt to the increased pulmonary artery pressure^4^. Mechanisms of RV maladaptation in PH are multifaceted. Emerging evidence suggests that lipid dysregulation involves in this process. Recent studies show that individuals with PH have lower high-density lipoprotein cholesterol (HDL-C), apolipoprotein A-I (ApoA-I), and low-density lipoprotein cholesterol (LDL-C), and higher triglyceride (TG) to HDL-C ratio than healthy control subjects^5–8^. In addition, lower HDL-C and LDL-C associate with disease severity and death; yet, the mechanisms remain unclear^5,6^.

Lipids and lipoproteins are established risk factors for left heart disease and atherosclerosis. Increased LDL-C and TG, and reduced HDL-C are associated with decreased systolic and/or diastolic left ventricle functions in patients with coronary heart disease^9^. Adult cardiomyocytes depend on lipids for energy production, accounting for approximately 60% of ATP production in the heart^10,11^. Interestingly, lipid utilization as energy source declines in the failing heart^10,11^. A recent study by Morisson et al^12^ showed an association between lipid metabolic markers and RV function and pressure, but the specific contribution of lipids to the development of RV failure in PH remains unknown. RV and LV have distinct hemodynamics and metabolisms, because of their distinctive roles within the cardiovascular system. Hence, assessing the contribution of circulating lipids to RV function in PH will help us gain more understanding of its mechanistic role in PH. We hypothesize that aberrant blood lipids relate to RV function and metabolism in PH.

## Methods

### Study design and population

This is a cross-sectional study of participants enrolled in The Pulmonary Arterial Hypertension Treatment with Carvedilol for Heart Failure (PAHTCH) clinical trial (NCT01586156) at baseline visit^13^. The study was approved by the Cleveland Clinic Foundation Institutional Review Board (number 11-1198) and all participants provided written informed consent. We included 42 subjects (n=12 control and n=30 PH). Participants were 18 to 65 years of age, World Health Organization (WHO) Groups 1, 3, and 4, and stable on PH therapies. Diagnosis of PH was confirmed by a historical right heart catheterization showing a mean pulmonary arterial pressure ≥ 25 mmHg and pulmonary vascular resistance above 3 Wood units according to the European Society of Cardiology/European Respiratory Society (ESC/ERS) guidelines at the time of study^14^. All clinical, laboratory, and imaging data were obtained at baseline visit of the PAHTCH study.

### Clinical data

Study participants provided data regarding demographics, body mass index (BMI), smoking status, co-morbidities, and statin, and fibrate therapy. We documented The New York Heart Association (NYHA) classification functional class at time of visit^15^. To better understand the association between lipid profile and disease severity, morbidity, and mortality, clinical outcomes of death or transplantation were collected from patients’ chart at the 5-year mark from the time of enrollment.

### Blood collection

All participants provided fasting blood samples. Plasma was isolated and stored at −80°C. The Cleveland Clinic Laboratory Diagnostic Core measured total cholesterol (TC), TG and lipoproteins (LDL-C and HDL-C) using the Cobas ce6000 with e601 module instrument (Roche Diagnostic, Indianapolis, IN). We assessed human apoA-I using the human apoA-I ELISA Pro kit (Mabtech Inc., Cincinnati, OH). NT-proBNP was measured by an electrochemiluminescence immunoassay using the Roche Cobas e411 analyzer as previously described^13^. Data less than 5 pg/mL are below the detectable level and are assigned an arbitrary value of 4.

### Echocardiogram

Study participants underwent a comprehensive transthoracic Doppler echocardiogram using Vivid 9 (GE Healthcare, Horten, Norway) and following the American Society of Echocardiography (ASE) guidelines. A single registered advanced cardiac sonographer (MP) performed this procedure to assess RV size, function, and right heart hemodynamics, utilizing standard two-dimensional (2D) imaging, m-Mode, tissue, and spectral Doppler as previously reported^13^. Cardiac sonographer measured the following parameters: right atrial (RA) dimensions, RA pressure (RAP), RV end-diastolic (RVED) and RV end-systolic (RVES) dimensions, RV wall thickness, RV systolic pressure (RVSP), RV fractional shortening percentage, RV tei-index, RV strain, and the tricuspid annular plane systolic excursion (TAPSE). For a better comprehensive assessment of the overall cardiac performance in PH, we also assessed LV systolic and diastolic function reflected by LV EF and LV tei.

### 2-[18F]fluoro-2-deoxy-D-glucose positron emission tomography (FDG-PET) uptake

Consented participants (healthy control, n=8; PH, n=30) underwent a FDG-PET scan after a minimum of 8 hours fasting period. Participants received an intravenous injection of 370 MBq (10 mCi) of [^18^F]-FDG. Following a 90-minute uptake period (average of 94±8 minutes, range = 77 to 119 minutes), images of PET/CT scanning (Biograph mCT, Siemens Molecular Imaging, Hoffman Estates, IL, USA) were collected. The scanner had a 22-cm axial field of view, which encompassed the heart and a significant portion of the lungs. For PET attenuation correction, a low-dose CT scan was performed, utilizing the following parameters: 120 kVp, 11 mAs, 4 mm slice thickness, and a pitch of 1.0.

A single-blinded radiologist (DRN) analyzed the fused PET/CT images to measure the standardized uptake value (SUV) in various cardiac structures, including the left ventricular (LV) free wall, interventricular septum, RV-free wall, and RA-free wall. Subsequently, the RV/LV SUV ratio was calculated.

### Statistical analysis

Quantitative variables were summarized with means ± standard deviation (SD), while categorical variables were summarized as frequency and percentage. We adopted log transformation when the data normality test failed. Group comparisons were done using Pearson’s χ2 or Fisher’s exact test for categorical variables and student t-test for continuous variables. Spearman’s rank correlation coefficient was calculated to study the association between continuous variables. All analyses were performed using JMP software (Cary, NC, USA). The level of statistical significance was set at *p*<0.05.

## Results

### Baseline and clinical characteristics

Participants with PH (N = 30) and healthy controls (N = 12) were enrolled in the study. Baseline characteristics of study participants have been reported previously^13,16^. PH patients had higher BMI compared to controls (PH, 29±6; controls, 25±4 kg/m^2^, *p*<0.01) and were more likely to have hypertension (PH, 33%; controls, 0%, *p*<0.04). There were no differences in reported dyslipidemia or use of statin or fibrate between PH and controls. Around 86.7% of PH patients were WHO group 1, 6.7% were WHO group 3, and 6.7% WHO group 4. The majority (63.3%) were NYHA class II (**Table 1**). The mean CAMPHOR total score was elevated indicating poor quality of life, this was reported in previous publication^16^. Previous publications reported echocardiogram and PET measurement parameters of the study cohort^13,16^. As expected participants with PH had worse RV function and increased RV glucose uptake by PET. After 5 years of enrollment, out of 30 PH subjects, one participant underwent lung transplantation, one died due to postpartum hemorrhage, one succumbed to hypoxemic respiratory failure due to ARDS secondary to acute pancreatitis, and one subject’s condition deteriorated prior to lung transplantation.

**Table 1.**
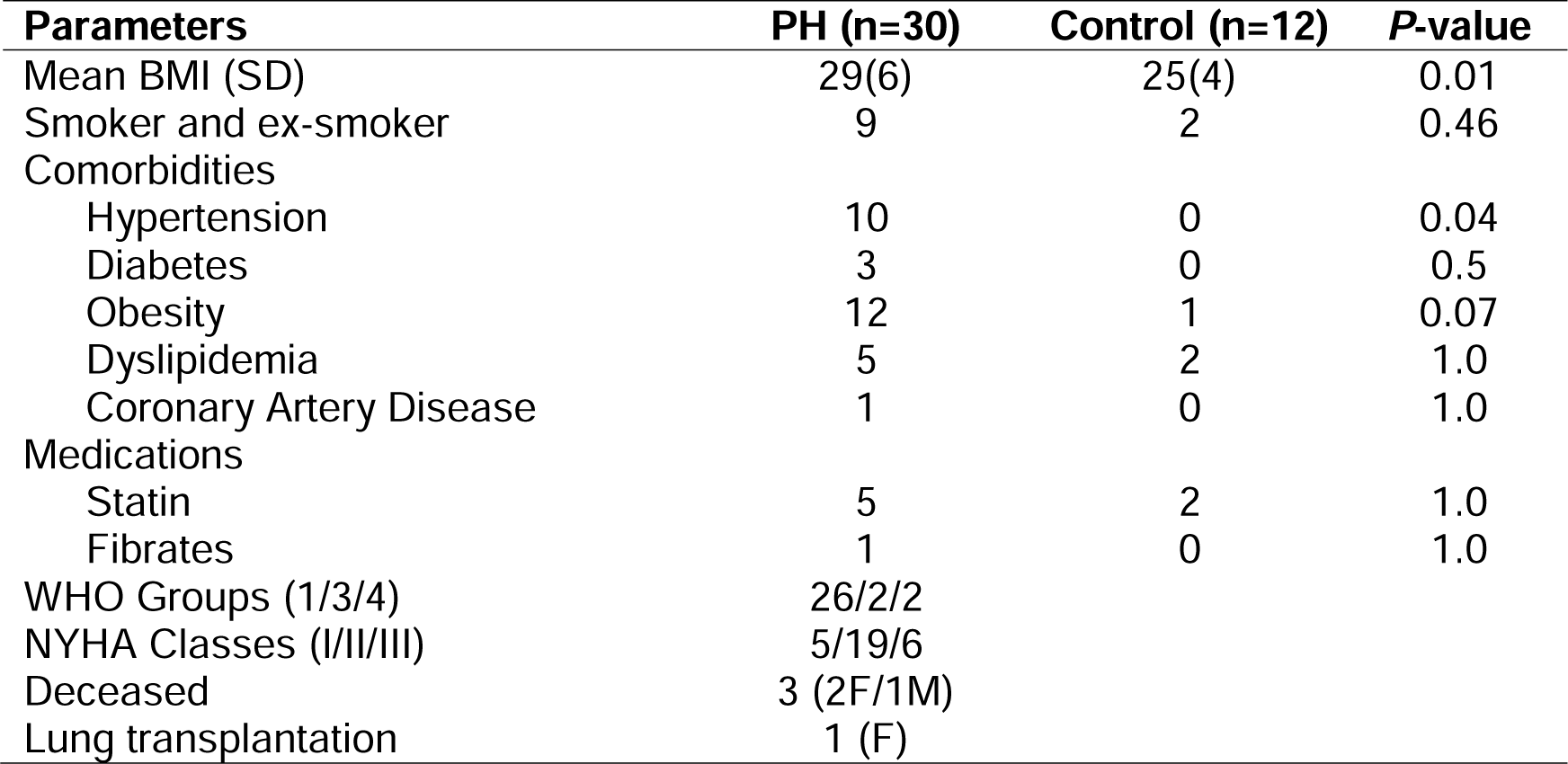
Clinical characteristics of PH patients vs. healthy controls.

| **Parameters** | **PH (n=30)** | **Control (n=12)** | **P-value** |
| --- | --- | --- | --- |
| Mean BMI (SD) | 29(6) | 25(4) | 0.01 |
| Smoker and ex-smoker | 9 | 2 | 0.46 |
| Comorbidities |  |  |  |
| Hypertension | 10 | 0 | 0.04 |
| Diabetes | 3 | 0 | 0.5 |
| Obesity | 12 | 1 | 0.07 |
| Dyslipidemia | 5 | 2 | 1.0 |
| Coronary Artery Disease | 1 | 0 | 1.0 |
| Medications |  |  |  |
| Statin | 5 | 2 | 1.0 |
| Fibrates | 1 | 0 | 1.0 |
| WHO Groups (1/3/4) | 26/2/2 |  |  |
| NYHA Classes (I/II/III) | 5/19/6 |  |  |
| Deceased | 3 (2F/1M) |  |  |
| Lung transplantation | 1 (F) |  |  |

### Lipid profile and plasma NT-proBNP levels

Compared to controls, subjects with PH had significantly lower HDL-C [PH, 41±12; controls, 56±16 mg/dL, *p*<0.01], higher TG to HDL-C ratio [PH, 3.6±3.1; controls 2.2±2.2, *p*<0.01] and lower apoA-I, the major apolipoprotein on HDL-C [PH, 1.7±0.5; controls 2.6±0.9 mg/dL, *p*=0.001]. TG tended to be higher in PH (PH, 125±66; controls 98±68 mg/dL, *p*=0.08). TC and LDL-C were similar between controls and PH (all *p*>0.1) (**Figure 1**).

**Figure 1.**
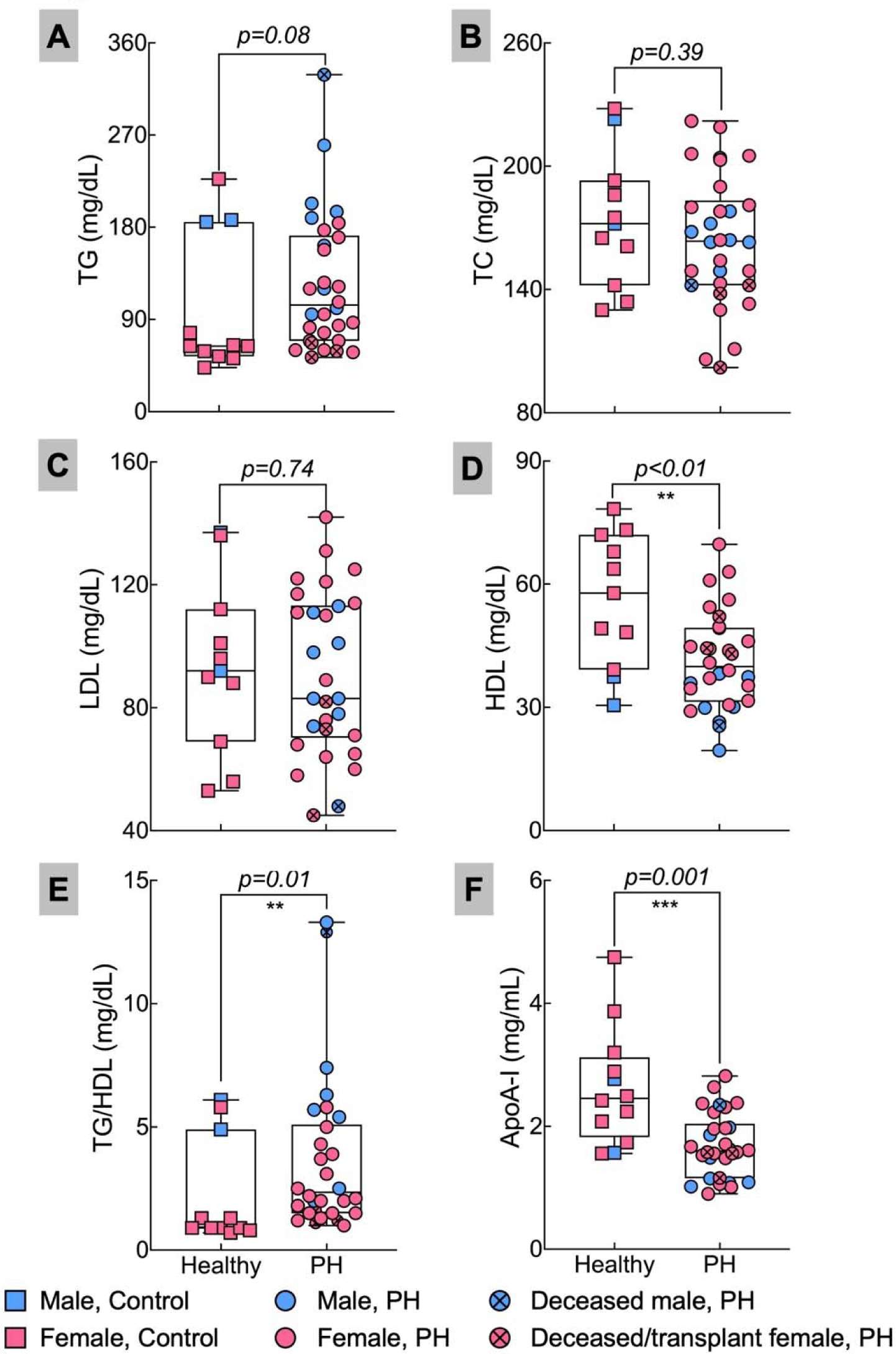
Differential plasma lipid distribution of TG (A), TC (B), LDL (C), HDL (D), TG/HDL ratio (E), and ApoA-I (F) in PH patients and healthy subjects. Plasma TG and ratio TG to HDL were not normally distributed according to Shapiro-Wilk normality test. Further analyses were performed with log-transformation.

### Association between lipids with right ventricle size, function, and glucose uptake

In PH patients, TC, LDL-C and TG significantly correlated with RVED and RVES area (**Figure 2**). TG inversely correlated with RVSP (*R*=−0.36, *p*=0.05). Both TG and TC correlated positively with percentage of RV fractional area change (*R*=0.51 and *R*=0.48, respectively, both at *p*<0.01) (**Figure 3**). TC negatively correlated with RV strain, while TG correlation with RV strain tended toward significance (**Figure 3**). TG was significantly associated with TAPSE (*R*=0.38, *p*=0.04). Lower TC correlated with higher NT-proBNP and higher RV glucose uptake, suggesting lower TC associates with reduced heart function and preferences for an increased RV glucose utilization in PH (**Figure 4**). TG negatively correlated with NT-proBNP (*R*=–0.62, *p*<0.01), but did not correlate with RV glucose uptake. HDL-C and apoA-I did not correlate with RV dimensions, function, and metabolism. Interestingly, lower TC and TG correlated with higher LV Tei-index in PH. A summary of our findings is reported in **Supplementary Table 1**.

**Figure 2.**
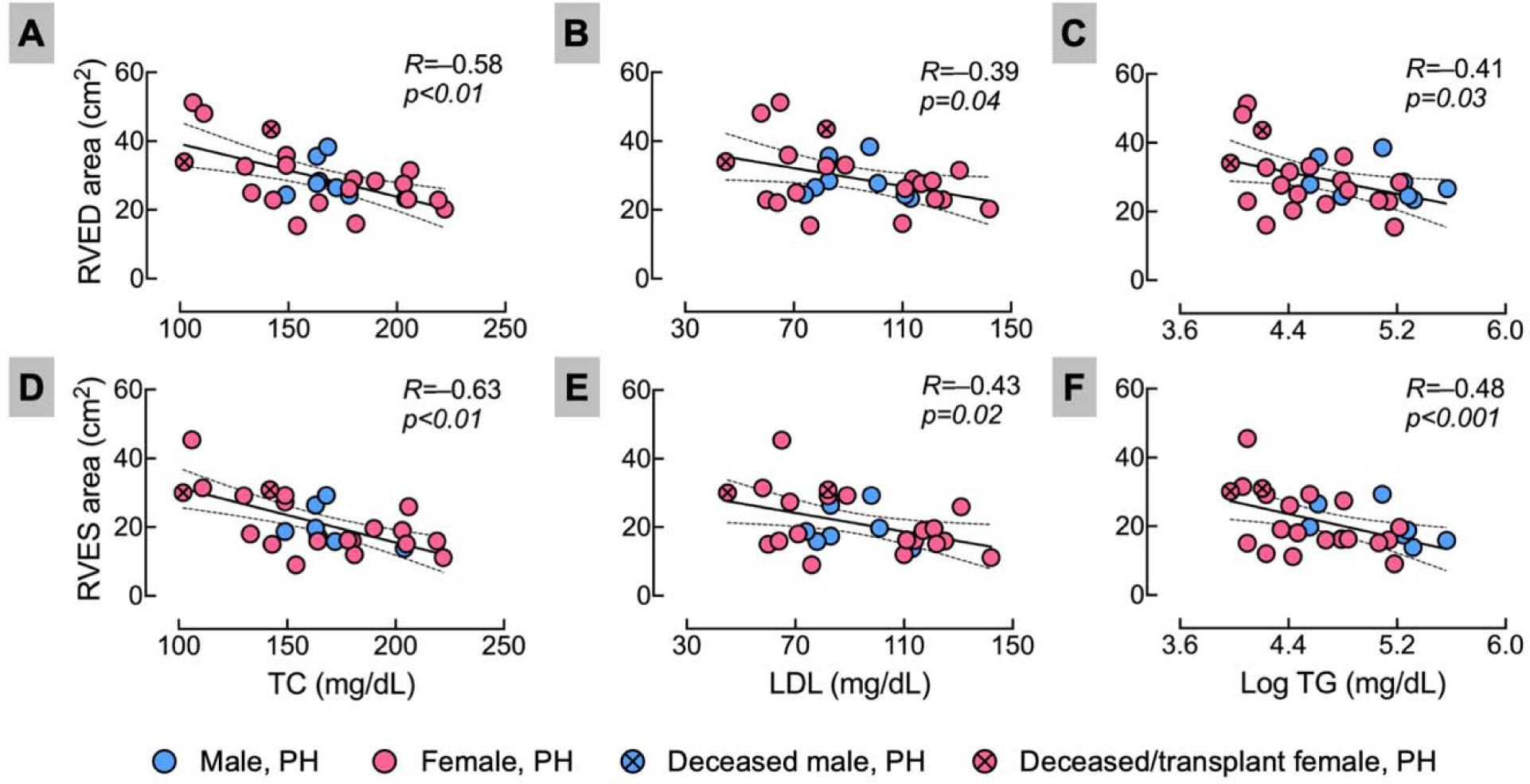
Lower TC, LDL and TG associated with RV dilation. Correlation of total cholesterol (TC), low density lipoprotein (LDL), triglyceride (TG) with RV dimension, measured as RV end-diastolic area (A, B, and C respectively) and RV end-systolic area (D, E, and F respectively) among PH patients. TG data were not normally distributed, data were log transformed.

**Figure 3.**
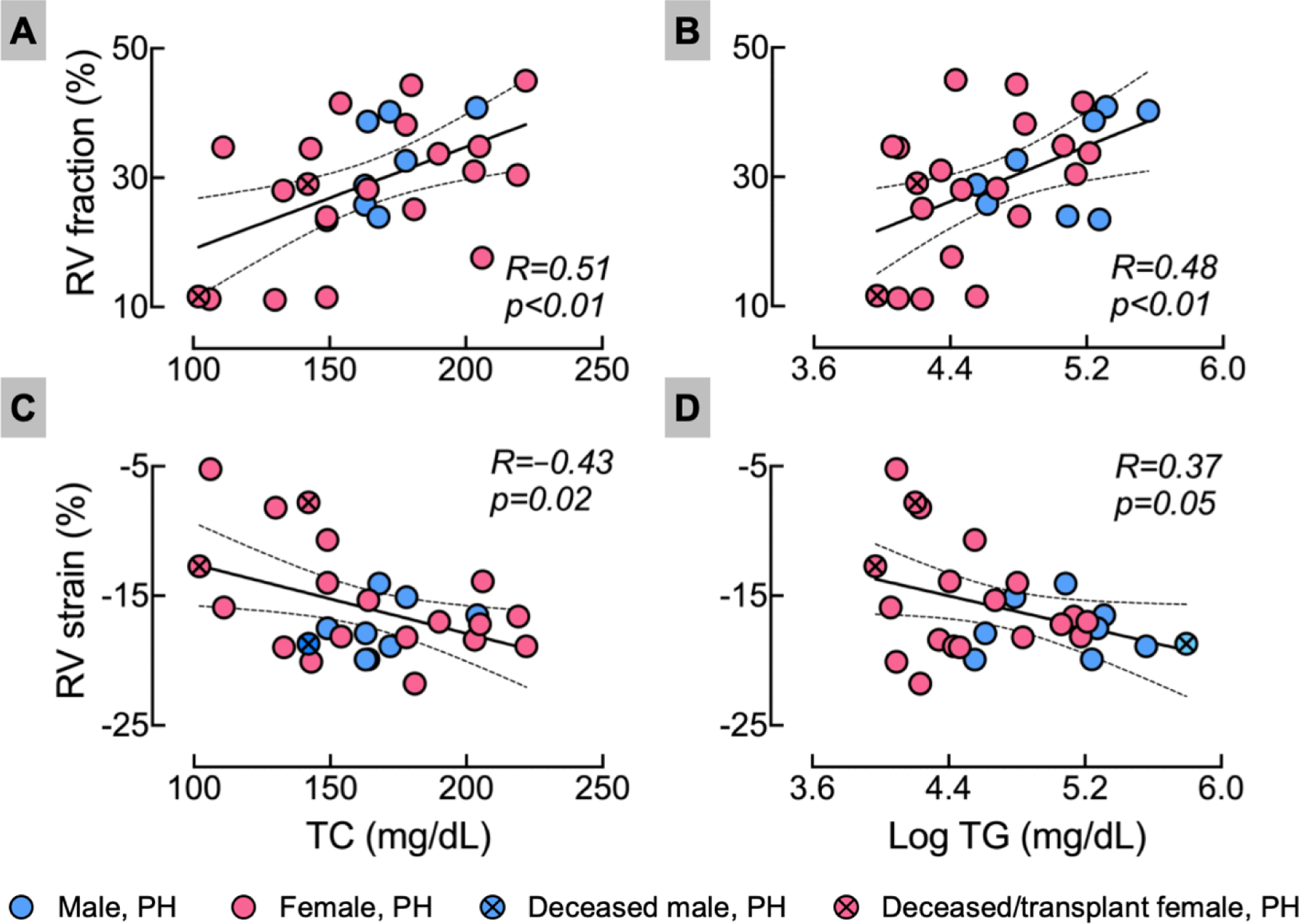
Lower blood TC and TG were linked to RV dysfunction measured as RV fraction and strain. Correlation of total cholesterol (TC) and triglyceride (TG) with RV fraction (A and B, respectively) and RV strain (C and D, respectively) in PH patients. TG data were not normally distributed, data were log transformed.

**Figure 4.**
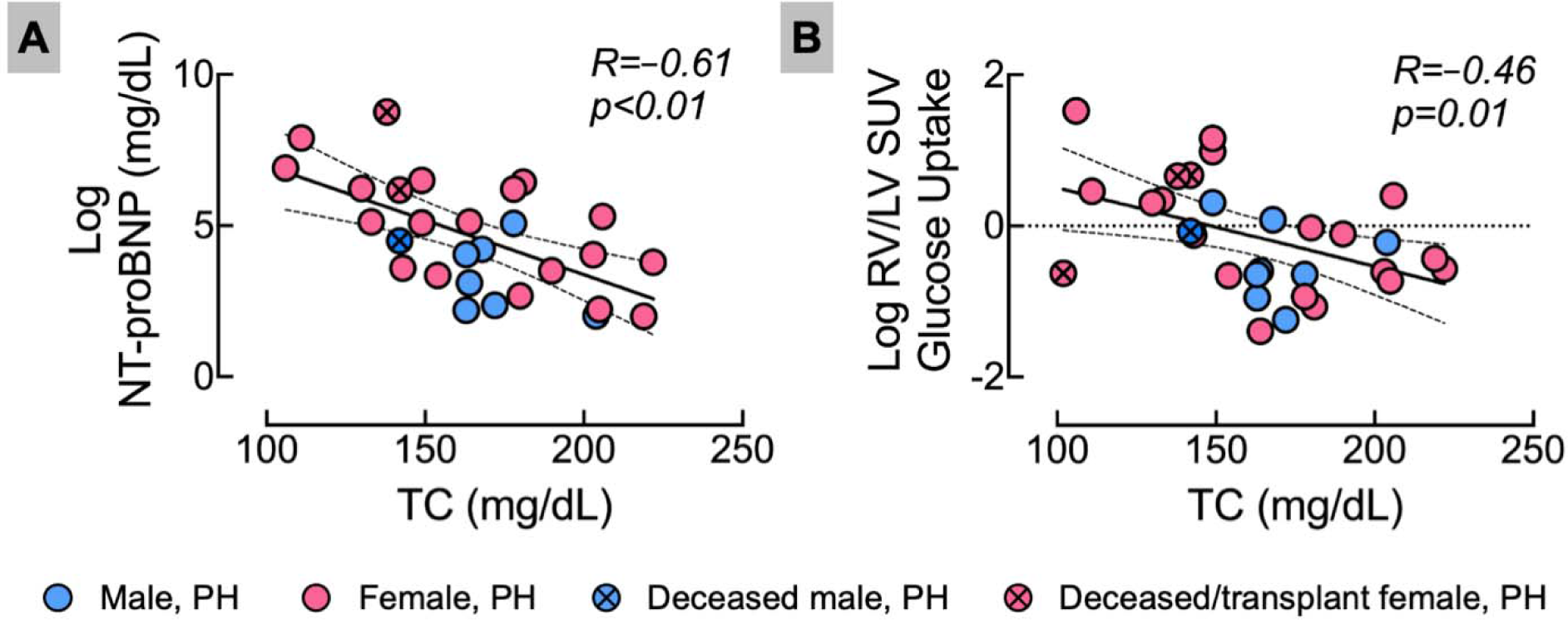
Lower blood TC was linked to higher plasma NT-proBNP and RV glucose uptake. Correlation of total cholesterol (TC) with NT-ProBNP (A) and RV/LV SUV (B) among patients with PH. Both NT-proBNP and RV/LV SUV data were not normally distributed, and correlation was performed in log-transformed values.

## Discussion

To the best of our knowledge, this is the first study that focuses on relationship between blood lipids and the RV function in PH patients. This study identified that lower blood lipids in PH, specifically TC and TG, are associated with (1) RV dilation, shown as higher RVED and RVES area, (2) RV dysfunction, reflected by lower RV fractional area change and higher RV strain, (3) aberrant RV metabolism, depicted by higher RV glucose uptake, and (4) higher NT-proBNP, marker of heart failure. Considering RV failure is the major determinant of PH severity and outcome, this association suggests that lipid metabolic pathway can be an important therapeutic target to improve RV function and clinical outcomes in PH.

In the current study, individuals with PH exhibit a distinctive plasma lipid distribution characterized by lower HDL-C, and apo-AI and higher TG to HDL-C ratio. This observation aligns with findings reported in previous studies^6–8,17,18^. A reduced level of HDL-C is an important biomarker and correlates with disease severity and mortality in PH^6,18–21^. Additionally, increased TG to HDL-C ratio associates with poor prognosis and is a risk factor for PH^18,22^. In patients with chronic thromboembolic PH, low apo-AI is a better biomarker for disease severity compared to HDL-C^8^. In line with current results, several studies show no alteration in TG level in PH^18,21^. While not confirmed in our findings, other research studies have documented reduced levels of LDL-C, and TC, demonstrating their prognostic role in PH^6,17,18^. These variations might be attributed to differences in participants’ demographic and clinical characteristics.

Several factors may account for differential circulating lipid distribution in PH. First, inflammation triggers alterations in lipid metabolism. Higher TG and lower HDL-C, leading to an elevated TG to HDL-C ratio. In severe cases, it may decrease LDL-C. This form of dyslipidemia is frequently encountered in various conditions, including rheumatoid arthritis, psoriasis, lupus and inflammatory bowel disease^23^. Furthermore, inflammation is a key element in the development and progression of PH. PH associates with numerous inflammatory disorders. Multiple studies have highlighted elevated levels of circulating cytokines, chemokines, tissue factors, and C-reactive proteins in patients with PH^24^. In addition, emerging data indicate correlations between low LDL-C and high TG to HDL-C ratio with systemic inflammation^5,7^. Bone morphogenetic protein type 2 receptor (BMPR2), is mutated in heritable PH and is reduced in other forms of PH^25^. BMPR2 may serve as a link between the roles of inflammation and lipid dysregulation in PH. On one hand, disruption of the BMPR2 signaling pathway results in increased inflammation, while on the other hand, BMPR2 mutation is associated with aberrant lipid metabolism^26,27^. Second, an abnormal lipid profile is frequently observed in cases of obesity and insulin resistance. In our study, PH group exhibits higher BMI compared to control group, that classified them clinically as overweight, a consistent finding supported by a substantial PH registry in the United States^28^. Additionally, previous studies have outlined disruptions in lipid homeostasis among PH individuals with insulin resistance^29,30^. Expanding on this, Zamanian et al, have demonstrated the TG to HDL-C ratio as a reliable surrogate for insulin resistance in PH, highlighting a valuable insight into aberrant metabolic dynamics in PH^22^. Third, it is crucial to recognize the role of the liver in both lipid/lipoprotein production and clearance. In the context of PH, RV failure and higher right-sided filling pressure can lead to congestive hepatopathy, ultimately progressing to liver fibrosis and cardiac cirrhosis. Recent studies indicate that increased serum bilirubin levels have been observed in 15 to 20% of patients with PH and are associated with high mortality risk. Particularly, direct bilirubin serves as a highly sensitive biomarker for venous congestion, response to treatment, and hemodynamic improvement in PH^31^. Moreover, chronic hepatic congestion contributes to the disruption of lipid metabolism in human and murine models by decreasing hepatic synthesis of lipoprotein, increasing hepatic clearance, and reducing intestinal absorption^31,32^. This shows the connection between how PH-induced changes in the heart, specifically RV dyfunction, influence lipid and lipoprotein metabolism in the liver. Liver function tests were not performed in this study. Nonetheless, the echocardiography findings of normal right atrial pressure in our PH population do not support hepatic congestion^13^.

The impact of dyslipidemia on LV function and failure is well-established^9^. However, the same observations are not clearly defined in RV. RV greatly differs from LV regarding structure, function, and metabolic pathways. The RV, characterized by its thin-walled, crescent-shaped structure, undergoes unique patterns of cardiac remodeling and bioenergetic shifts in response to similar pathological stimuli when compared to the LV^33^. Our study supports existing evidence, demonstrating an inverse relationship between RV function and TC, LDL-C, and TG. Notably, our findings highlight a significant correlation between TC with various RV function parameters, despite no significant differences from the control group. Aligning with our observations, a study involving 442 patients with congestive heart failure demonstrates an increased in RVED size correlates with lower levels of TC, TG, HDL-C and LDL-C^34^. While, in the Multi-Ethnic Study of Atherosclerosis of 4204 participants, RVED volume positively associates with HDL-C and inversely with TC^35^. These differences can be attributed to the differences in patient demographics and heart failure etiologies in these two study cohorts. Regarding RV function parameters reflected by RV fraction, strain, and TAPSE, our findings resonate with previous reports. In a study of 69 patients with idiopathic PH, HDL-C and TC positively correlates with TAPSE and RV fraction^17^. Additionally, in a retrospective cohort of 37203 participants referred for transthoracic echocardiography, a categorical increase in RVSP is associated with lower LDL and HDL (*p*<0.001)^12^. Furthermore, a categorical decrease in TAPSE was linked to a higher TG:HDL ratio and lower HDL and LDL (p<0.001)^12^. In contrast, in the Multi-Ethnic Study of Atherosclerosis of 3,032 participants, a high TG to HDL-C ratio associates with a lower TAPSE, RV fraction, and RV strain, along with higher odds of RV diastolic dysfunction (OR, 1.19; 95% CI, 1.03 to 1.39; *p*LJ<LJ0.05)^22^. Inconsistency in these previous studies may be attributed to differences in the etiology of heart failure and/or population differences.

Metabolic derangements of cardiac tissue, including lipid metabolism, are evident in PH population^36^. A normal adult heart utilizes enormous amount of ATP to sustain the contractile function. The major source for ATP production in the adult heart is mitochondrial oxidative phosphorylation, providing 90% of total myocardial ATP requirement, and lipid is the fuel preference for cardiomyocytes, contributing up to 60% of total energy produced^11^. Energy metabolism is altered in heart failure, described as (1) less ATP content in the failing heart compared with a normal heart, and (2) preferences for glucose metabolism via glycolysis as the source of energy in response to reduction in lipid oxidation^11^. Similarly, PH presents with cardiolipotoxicity phenotype^27,29,37^ as evidenced by (1) changes in energy metabolism with reduced mitochondrial fat oxidation^38–40^, (2) preferences for higher RV glucose uptake in PH associate with poorer outcome, higher death or transplantation^41^, and (3) lipid deposition in cardiac tissue^27,42^. The current report adds evidence that shifting toward glucose metabolism in the RV of PH patients associates with lower TC, representing the total circulating lipoproteins. The source of fatty acids delivered to the heart is derived from fatty acids bound to lipoproteins or albumin in the blood, suggesting there may be an association between lipoproteins and cardiac energy metabolism. However, the role of circulating lipids and lipoproteins on the abnormality in cardiac lipid metabolism in PH is not well-established and warrants further investigations.

Current findings regarding the association of TC with RV function are aligned with the concept that a lower level of TC links to a poorer prognosis and adverse clinical outcomes. This phenomenon has been documented in various populations with chronic diseases, including in patients with heart failure^43^, in aging population^44^ and several other groups. The restoration of TC levels to normal ranges can potentially alleviate the negative impacts linked to systemic inflammation and malnutrition, thereby leading to improved patient outcomes.

Our study has limitations, which may influence the strength and clinical relevance of the deductions drawn from our results. First, this is a cross-sectional study, therefore, establishing causal or temporal relationships between lipids and RV function in PH is challenging. Future prospective studies are warranted to gain a comprehensive understanding of whether circulating lipid levels associate with the progression of RV dysfunction over the course of the disease. Second, this study is single-center in nature and has a small sample size, which limits the generalizability of current findings. Multi-centered studies with larger sample sizes are recommended to validate our results. Third, potential confounding factors, such as medication use, comorbidity, dietary intake, and lifestyle factors, may influence our findings, particularly regarding lipid distribution. Fourth, we could not assess the involvement of liver in lipid dysregulation in the context of PH due to the absence of specific liver function test parameters. Additionally, liver function test parameters were not collected from control subjects, further limiting our assessment.

In summary, PH patients have a distinctive plasma lipid distribution pattern that is associated with worse RV function. Thus, understanding both circulating and cellular lipid metabolism in relation to RV metabolism and function will provide valuable insights into mechanisms and potential targets for treatments.

## Data Availability

All data produced in the present study are available upon reasonable request to the authors.

## Acknowledgements

We thank all our research participants, and the Clinical Research Unit staffs at the Cleveland Clinic (Cleveland, OH) for assistance with participant screening and clinical visits. We thank Alan Pratt from the Cleveland Clinic Laboratory Diagnostic Core for blood lipid measurements. We thank Allison Janocha and Michael Novotny for language editing. We acknowledge the use of Grammarly Inc. for proofreading.

## Funding Supports

This research was supported in part by the following grants: R01HL115008 (SCE), R01HL060917 (SCE) and 4UL1TR002548 (CTSC, AM - pilot grant)

## Authors contributions

Conceptualization – BF, SAAC, SF, SCE and AM

Methodology – BF, SAAC, FPD, DRN, FPD, SF, SCE and AM

Formal analysis – BF, SAAC, SF and AM.

Investigation – BF, LP, SAAC, JS, MMP, DRN, FPD, SF, and AM

Resources – LP, SAAC, and SCE

Data curation – BF, LP, SAAC, JS, MMP, DRN, FPD, SF and AM

Writing – Original Draft: BF and AM

Writing – Review and Editing: BF, WHWT, FPD, SF, SCE and AM

Supervision – SF, SCE and AM

Funding Acquisition – SCE and AM

## List of Abbreviations

ApoA-I: Apolipoprotein A-I
ASE: American Society of Echocardiography
ATS: American Thoracic Society
CAMPHOR: Cambridge Pulmonary Hypertension Outcome Review
FDG: 2-[18F] fluoro-2-deoxy-D-glucose
HDL-C: High-Density Lipoprotein Cholesterol
LDL-C: Low-Density Lipoprotein Cholesterol
LV: Left Ventricle
NT-proBNP: N-terminal pro-B type Natriuretic Peptide
NYHA: New York Heart Association
PAHTCH: Pulmonary Arterial Hypertension Treatment with Carvedilol for Heart Failure
PET/CT: Positron emission tomography / computed tomography
PH: Pulmonary Hypertension
RA: Right Atrium
RAP: Right Atrial Pressure
RV: Right Ventricle
RVED: Right Ventricle End-Diastolic
RVES: Right Ventricle End-Systolic
RVSP: Right Ventricle Systolic Pressure
SUV: Standardized Uptake Value
TAPSE: Tricuspid Annular Plane Systolic Excursion
TC: Total Cholesterol
TG: Total Triglycerides
VLDL-C: Very low-density lipoprotein cholesterol
WHO: World Health Organization

---

**Supplementary Table 1:** Correlation of lipids and lipoproteins with RV dimension, function, and metabolism and plasma Pro-BNP in PH patients only.

|  | TG (mg/dL) |  | TC (mg/dL) |  | LDL (mg/dL) |  | HDL (mg/dL) |  | apoA-I (mg/mL) |  |
| --- | --- | --- | --- | --- | --- | --- | --- | --- | --- | --- |
|  | R | p | R | p | R | p | R | p | R | p |
| RVED area (cm<sup>2</sup>) | **-0.41** | **0.03** | **-0.58** | **<0.01** | **-0.39** | **0.04** | -0.24 | 0.21 | -0.20 | 0.31 |
| RVES area (cm<sup>2</sup>) | **-0.48** | **<0.01** | **-0.63** | **<0.01** | **-0.43** | **0.02** | -0.19 | 0.34 | -0.18 | 0.37 |
| RVSP (mmHg) | **-0.36** | **0.05** | -0.28 | 0.13 | -0.24 | 0.21 | 0.22 | 0.25 | 0.04 | 0.81 |
| RV thickness (cm) | -0.19 | 0.31 | -0.34 | 0.07 | -0.21 | 0.26 | -0.13 | 0.48 | 0.20 | 0.29 |
| RV fraction (%) | **0.48** | **<0.01** | **0.51** | **<0.01** | 0.36 | 0.06 | -0.05 | 0.79 | 0.04 | 0.82 |
| RV strain (%) | **-0.37** | **0.05** | **-0.43** | **0.02** | -0.21 | 0.29 | -0.14 | 0.46 | -0.24 | 0.22 |
| RV Tei-index | -0.24 | 0.19 | -0.26 | 0.17 | -0.12 | 0.54 | -0.07 | 0.72 | -0.25 | 0.19 |
| TAPSE | **0.38** | **0.04** | 0.23 | 0.23 | 0.11 | 0.55 | -0.23 | 0.22 | 0.07 | 0.69 |
| RV/LV SUV | -0.30 | 0.10 | **-0.46** | **0.01** | -0.25 | 0.19 | -0.18 | 0.34 | -0.07 | 0.71 |
| NT-proBNP | **-0.62** | **<0.01** | **-0.61** | **<0.01** | -0.36 | 0.05 | -0.17 | 0.38 | -0.16 | 0.40 |
| LV EF (%) | -0.08 | 0.67 | -0.08 | 0.69 | -0.01 | 0.94 | -0.01 | 0.97 | -0.12 | 0.53 |
| LV Tei-index | **-0.55** | **<0.01** | **-0.43** | **0.02** | -0.23 | 0.21 | 0.14 | 0.47 | -0.13 | 0.51 |

